# Understanding the influences on the design and delivery of an integrated child health and social care service in underserved communities in the UK: A qualitative exploration using the SELFIE framework

**DOI:** 10.1101/2024.10.03.24314613

**Authors:** I Litchfield, L Harper, S Abbas, F Dutton, M Melyda, C Wolhuter, C Bird

## Abstract

**Background:** The UK’s National Health Service has provided funds for developing localized services integrating health and social care intended to address the health inequities prevalent in children and young people living in marginalized communities. However, little is understood of the factors that influence their design and delivery, nor which combined health and social care models are most effective.

**Objective:** To use evidence drawn from staff delivering a collocated integrated health and social support service to inform future integrated care offers.

**Methods:** A qualitative exploration of staff experience using a directed content analysis to populate and present the results within the Sustainable integrated chronic care model for multi-morbidity: delivery, financing, and performance (SELFIE) framework. The analysis presented here focusses on the domain of *Service delivery*, predominantly relating to the content and access of care.

**Results:** A total of 14 staff were interviewed: clinicians from primary and secondary care, social care providers, local voluntary groups, and school-based family mentors. Participants described at the *Micro-* level how the service increased engagement of families and facilitated referral to social support and preventative care; at a *Meso-* level the benefits of collocation, collaborative working, and community outreach were described. Finally at the *Macro* level, improvements to the access and availability of appropriate care were observed.

**Conclusions:** The pilot appeared to deliver multiple benefits for both patients and staff and the broader health economy particularly through collocating health care and social support. However, sustainable integrated care requires greater institutional commitment and leadership.

**Research in Context:** *What is already known abou t the topic?:* In the UK, the National Health Service England has been reorganised to facilitate closer collaboration between health and social care organisations. This includes prioritizing and funding localized services that integrate multiple strands of clinical, preventative and social care. Despite these policy intentions there are few lasting examples that have produced practical learning of which the ‘Sparkbrook Children’s Zone’ is one.

*What does this study add to the literature?:* Participants described how school outreach, the multidisciplinary team, and extended consultation times increased engagement of underserved families. The collocation of health and social support, both improved referral rates and allowed for more personalised care. Despite the positive experience of staff and patients and the improvement in access and availability of health and social care there was a perceived lack of support at a system level.

*What are the policy implications?:* Carefully fostered links with local schools meant the service was better able to identify and reach vulnerable families earlier and helped address issues of trust around mainstream healthcare that can exist in underserved populations. The collocation of social support allowed for direct same-visit referrals between services and the chance to address underlying issues.

## Background

The challenges to the health and well-being of children and young people in marginalized populations include an increased prevalence of chronic conditions, obesity, and mental ill health [1–3]. They are exacerbated by a range of socio-economic and cultural pressures that inhibit and utilisation of primary or preventative health care services; social determinants of health (SDoH) that include income, housing, and food insecurity [4–7]. Health institutions in many high income countries are recognising that a more holistic approach is needed to address these [8, 9]. To this end policymakers and commissioners in multiple health systems are encouraging collaboration between health services, social care providers, local authorities, voluntary, community and faith sector (VCFS) groups, and other agencies to improve health and reduce health inequalities [10–12]. This has led to the introduction of new models of integrated health and social care emerging in Australia [13], North America [14, 15], and across Europe [16].

In the UK, the National Health Service England (NHSE) has been reorganised under the Health and Care Act 2022 to facilitate closer collaboration between health and social care organisations. [17, 18]. This includes prioritizing and funding localized service delivery that integrates multiple strands of health and social care [10, 19–25]. One such example that combines General Practitioners (GPs), family support workers, mental health outreach, and paediatricians is the Sparkbrook Children’s Zone (SCZ) [26]. Its intention is to treat and manage acute and chronic health care alongside providing the necessary social support for CYP and their families [27, 28]. The different elements of the SCZ summarising the visible elements involving the contact between patients and providers and the invisible processes and infrastructure that support its delivery are summarized in Figure 1.

**Figure 1:**
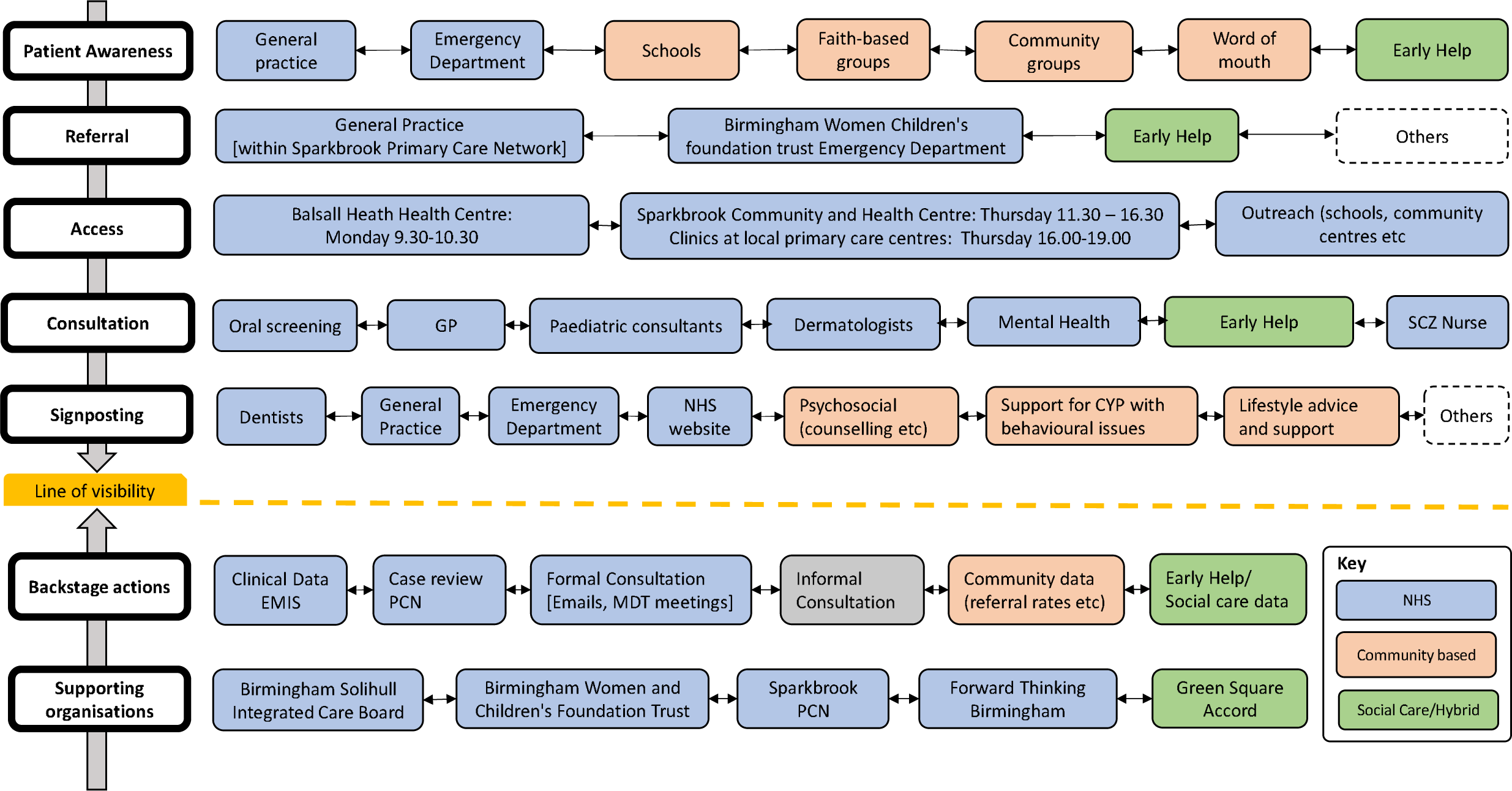
Service blueprint outlining Sparkbrook Children’s Zone integrated service

Despite the introduction of pilot programmes such as the SCZ little is understood of the factors that influence their design and delivery, nor which health and social care models are most effective [29–33]. The work presented here presents an in-depth exploration of the factors that shaped the SCZ, describing the experiences of the range of staff that deliver the service. The findings are presented within an *a priori* framework designed to examine and support integrated care [34], offering structured insight into the facilitators, barriers, and benefits of delivering integrated place-based health and social care in the UK.

## Methods

### Study design

The work consists of a qualitative exploration of staff perspectives using data gathered from a series of semi-structured interviews and analysed using the “*Sustainable intEgrated chronic care modeLs for multi-morbidity: delivery, Financing, and performance”* (the SELFIE framework) [34]. The framework consists of a number of coordination concepts from micro-through to macro-levels incorporated within six domains informed by the World Health Organisation’s interpretation of healthcare systems (see Supplementary File 1) [35]. The work presented here specifically explores the factors affecting the delivery of the service including availability and access and focuses on the domain of *Service delivery*, relating to the (see Table 2) [34]. Our sister paper uses the remaining domains of the SELFIE framework to explore the infrastructural and organisational factors underpinning the delivery of the SCZ.

### Population/recruitment

The SCZ is based in Sparkbrook & Balsall Heath East ward in Birmingham a large and diverse city in the UK’s midlands. It is the second most populous ward in the city, has the second highest level of deprivation and a superdiverse, young population with high rates of unemployment and some of the highest levels of infant mortality in England. It is also disproportionately affected by childhood obesity, child criminal and sexual exploitation, poor housing, chronic disease, and high levels of universal needs around housing, food, clothing, sanitary products, and essential supplies [36].

All staff involved in developing, managing and delivering the SCZ were eligible for inclusion. They were approached by [1st author] [3rd author] and [7th author], in-person or via email: all were supplied with a participant information sheet, and the opportunity to ask questions of their participation; ultimately providing informed consent before the interview commenced. We aimed to carry out interviews with 5-6 service providers from each organisation (including service leads, those actively delivering the service and administrative/support staff) to reach a total of 25 interviews sufficient to provide a rich and representative data set [37].

### Data Collection

Semi-structured interviews were conducted online (via Teams or Zoom), face-to-face in a room at the clinic, or via telephone by [First author] and [Third author] experienced qualitative researchers unknown to participants. They are experienced qualitative researchers that used a topic guide informed by the existing literature and covering a range of themes including experiences of engaging with the local Integrated Care System, barriers and facilitators to delivering the SCZ, and reflections on its future development. Digital audio recordings were transcribed verbatim by an approved third-party transcription service and the data were managed using nVivo vs12.

### Data analysis

Two authors [first author] and [third author] independently coded each transcript fitting the data within each of the relevant themes of the SELFIE framework using a directed content analysis [38] that allowed the identification and inclusion of emerging domains, constructs or sub-constructs [39]. Any differences in coding were discussed between the two authors and a consensus arrived at. The final allocation of the data within the coding framework was agreed by all authors. The work presented here explores the data through the SELFIE domain of *Service Delivery*.

## Results (1769)

### Characteristics of participants

We interviewed 14 participants over 13 interviews (two participants were interviewed at the same time). The interview lasted between 18 and 70 minutes. Of the 14 participants five were from primary care, three secondary care, two from social support, one that worked in local education, and one for a children’s charity.

**Table 1.** Characteristics of participants

| ***Participant ID*** | ***Sector*** | ***Role/Job Title*** |
| --- | --- | --- |
| 01 | Secondary care | Senior staff |
| 02 | Education | Family mentor |
| 03 | Primary Care | GP |
| 04 | Secondary Care | Consultant |
| 05 | Secondary Care | Consultant |
| 06 | Primary Care | GP |
| 07 | Social support | Family Support |
| 08 | Integrated care system | Operations manager |
| 09 | Primary care | Health Care Associate |
| 10 | Primary care | Health Care Associate |
| 11 | Children's charity | Service lead |
| 12 | Primary care | GP |
| 13 | Social support | Project manager |
| 14 | Social support | Service lead |

### Qualitative data

Below we present our findings within each of the relevant constructs within the Service Delivery domain at Micro -, Meso-, and Macro-levels. They are described alongside exemplar quotes identified by participant ID, and Job role. These findings are summarised in Table 2.

**Table 2.**
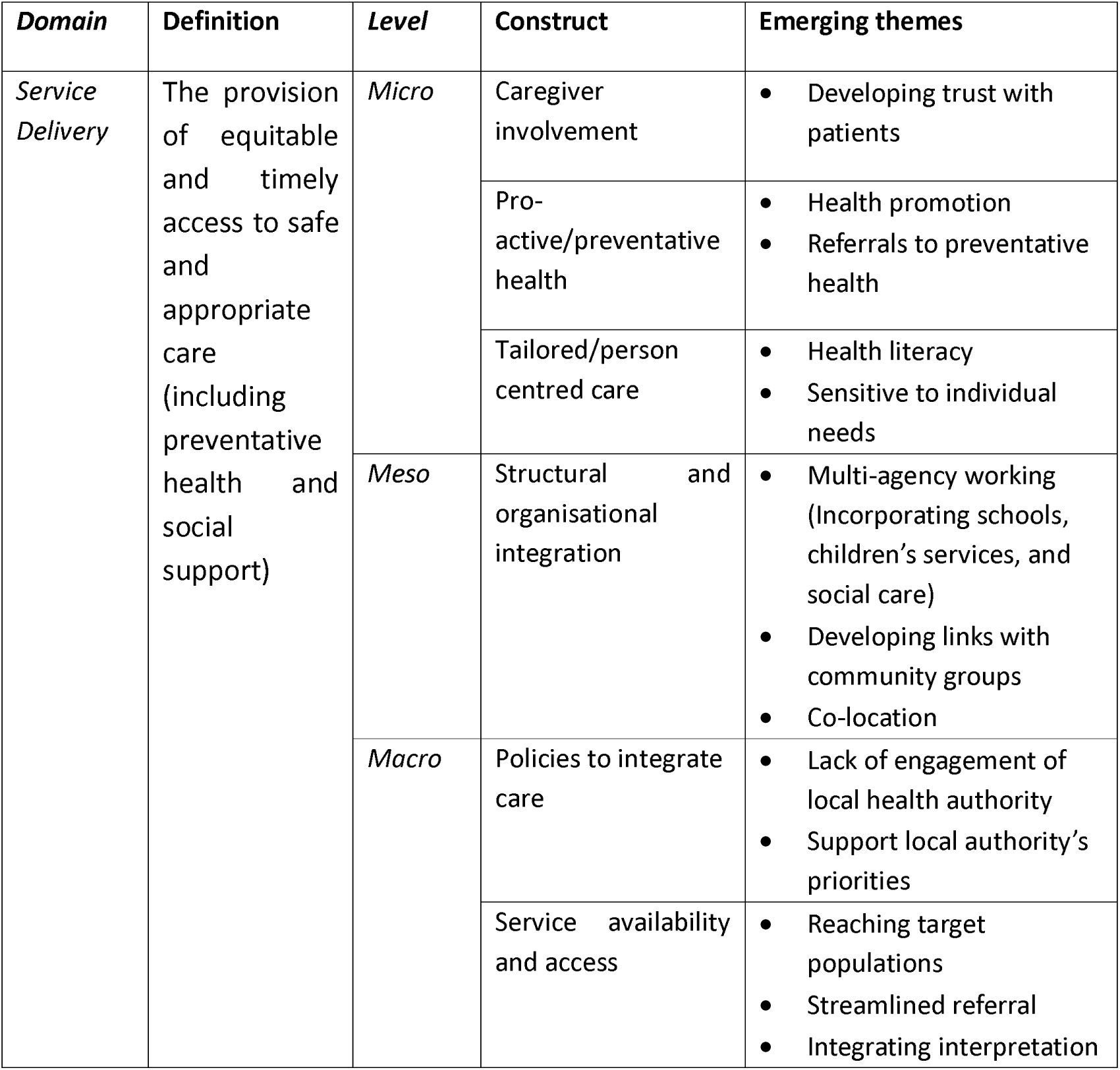
Summary of SELFIE informed analytical framework and emerging themes [34]

### Micro-

#### Caregiver involvement

Clinicians and social support providers understood the importance of fostering trust and developing a collaborative alliance with parents. This included the understanding that (frequently attending) parents often needed time to talk about the wider challenges they face:

> “…they come to Early Help, we have a long conversation with them, and they go away feeling better because they actually feel like they’ve had their problems and issues listened to…”
>
> P07, Social Support, Family Support

#### Pro-active/Preventative health

Clinicians in the SCZ seized the opportunity to engage parents in health promotion by using the responses to a wider set of contextual questions to link patients directly with locally available preventative health offers:

> “because the clinicians are looking more broadly at health and wellbeing of that child - in its place, in the family, and in the community… the family is learning things that maybe it didn’t even know were available…[SCZ] is able to say, “Oh, this is something that you could do now, it could help your child lose weight, this could help your child brush his teeth better…”
>
> P01, Secondary Care, Consultant

#### Tailored and personalised care

The way in which the SCZ was designed meant that the social support practitioners were able to reach families that had been referred for clinical reasons but then address the specific needs of individual families:

> “…we can also educate our parents of how to help themselves, so they become more resilient, and have an understanding of how best to parent their child, care for their child, meet their child’s needs, where to gain the support when it is needed, and break down those barriers as well.”
>
> P07, Social Support, Family Support

### Meso-

#### Structural and organisational integration

There was an awareness amongst all of those involved that offering joined-up health and social support allowed them to more holistically address the needs of the patients:

> “…families present to GPs at the children’s zone, but often they’re coming to have symptoms treated, where actually the underlying cause isn’t always medical, sometimes it’s more of an Early Help need…if we can address both of those things, treat the symptoms and hopefully the cause, it will result in less presentations in the future.”
>
> P07, Social Support, Family Support

A significant part of the SCZ’s attempt to reach target populations were local schools. This included staff from the SCZ both proactively arranging sessions on a predesignated issue and responding to requests from schools to meet families they were concerned about:

> “We will contact a school, and then we’ll say, “Can we come in and talk about picky eating?” And then we’ll say… or diabetes, or some kind of health thing, and then they’ll pick their families, so they know, or more successfully schools will then say, “We’re seeing there’s a really… there’s low attendance with some of our children, and we don’t really know why, they say it’s health related but we don’t really know why, can you come in?”
>
> P13, Social Support, Project Manager

Not all of the organisational integration was formal and predetermined, the location of the service at the heart of target communities also helped create organic and productive partnerships with local groups because of their physical presence in the space:

> “…we’ve done work with the youth centre, which is about two/three minutes’ walk away, and so we’ve been able to share the stuff that we know about the [SCZ] with the youth centre, and it just all starts to make… create a professional network. It’s not formal, it doesn’t have to meet round a Teams meeting, but it’s just being in a community and in a space, and learning about what’s on offer…”
>
> P11, Children’s Charity, Service Lead

The benefits of collocating organisations and members of the multi-disciplinary team were reported by patients who appreciated the ability of the SCZ to address multiple issues in one visit, for example the opportunity to visit PAUSE a children’s health and well-being counselling service[40]:

> “We had [secondary care consultant] … walk a family over to the drop-in [PAUSE], and they were given support there and then, and mum and child were absolutely amazed that they could from having a meeting that was scheduled that week with a GP to having attended that, and within - I think it was an hour - having had the intervention, that the GP has signposted to. I think that slightly blew mum’s mind…being like ‘That doesn’t normally happen! How is this…?”
>
> P11, Children’s Charity, Service Lead

The collocation also enabled staff to deliver better care by being more aware of what each service provided and being able to consult with colleagues from other settings in-person:

> “I get to discuss them [the patients] with the consultant who’s just next door …we don’t really get that opportunity as GPs, we sit in our own room, doors are closed, we have the patients come in, we want to get advice we speak to a paediatrician over the phone. It’s not the same as actually discussing with someone who’s right there physically, who can actually walk into my room and see my patient in front of me. It’s completely different, it’s an invaluable experience… you can’t replicate that experience over a phone call”
>
> P12, Primary Care, GP

### Macro-

#### Policies to integrate services

The original idea for the SCZ came from emergency department clinicians at the local children’s hospital attempting to address the disproportionate numbers of parents presenting from the most deprived wards in the city. Despite the SCZ meeting key priorities of the local (Birmingham and Solihull) Integrated Care System, responsibility for the ongoing management of the service, including securing funding remained with the clinicians that were also those delivering frontline care:

> “…I feel like the leadership for this has been largely on the backs of the doctors running the service…this is a service that reduces health inequalities, this is the priority for the ICS…the ICS has not enabled them in the way that if this was a real priority you’d see more leadership I think from within the ICS for developing this.”
>
> P01, Secondary Care, Consultant

It was understood that the SCZ would also help inform the local authority’s approach to developing cross-sector support for underserved families:

> “In two or three ways this model is essentially very good. One is the [council] is thinking about family hubs so this model is really working on family hub level, and it is doing exactly what family hubs are supposed to do, bring a range of services together in one area. So this model could be a good pilot, an idea for a family hub, how health and non-health actually works together.”
>
> P03, Primary Care, General Practitioner

#### Service availability and access

Local schools were understood to be a valuable means of accessing local families that might otherwise fail to engage with mainstream health care [41]:

> “…at our school we’ve got a lot of Arabic Yemeni, we’ve got a recent migration of parents from Somalia and Nigeria. So, it’s a lot about maybe targeted groups such as parents who may not have English as their first language and may be hesitant about going to the doctors…these initiatives… can really help to reach out to our families who may be more deprived or isolated within society.”
>
> P02, Education, Family Mentor

The further engagement of local underserved families was supported by holding informal, group consultations in the familiar, non-clinical environment of their child’s school:

> “The reason why [parents attend] is because… it’s in a relaxed environment, it was a coffee morning, so it wasn’t a doctor’s surgery, it wasn’t an intimidating environment, it was a safe known environment to parents, and also it was done in a bit of a group discussion. … it was nice the fact that they were actually able to speak to the doctor and if they didn’t understand anything Doctor [GP’s name] was brilliant at explaining - for example there was something about birth marks…”
>
> P02, Education, Family Mentor

The colocation of social support and clinical care meant parents could not only access the clinical care for which they were referred but also social support without the associated stigma:

> “…and that is critical for Sparkbrook Children’s Zone, because there’s no shame in taking your children to the doctor. That is… going to the doctor is a safe and legitimate activity. So, I think that’s even though not everybody going to the doctor needs support, there is a way of… that is one of the benefits.”
>
> P14, Social support, Service Lead

##### Streamlined referral processes

The benefits of being able to refer instantaneously between the various elements of the service made it easier to access for all families, but particularly those with competing priorities that might otherwise be unable or unwilling to commit to additional appointments and further visits:

> “I don’t need to do a long several page referral into Early Help. As a doctor I’m saying, “There you go, you can just go next door”…that’s a service that the family would have been able to access by picking up the phone… I think we know that families’ motivation and resilience can be so low when they are struggling with lots and lots of different things in your life… that they’re not going to pick up the phone.”
>
> P04, Secondary care, Consultant

### Interpreters as a routine part of the service

Staff valued the ability of the SCZ team to provide the majority of parents and patients with interpreters and the opportunity to ensure that they understood what was happening as they were referred between services within the SCZ:

> “There are obviously some language needs, which we have met through our telephone interpreting service, which has been really good…Some of the [referring] GP practices have been really good at booking in advance interpreters for patients and their parents and carers so that they’ve got no language barrier when they come to clinic, which is especially excellent. It doesn’t always happen, but we’ve always got a service that will be able to translate, which is really nice.”
>
> P05, Secondary care, Consultant

## Discussion

### General findings

The combination of primary, secondary care, social support, allied children’s services and preventative health offered by the Sparkbrook Children’s Zone is designed to provide holistic care capable of addressing the social determinants of health. Using the Service Delivery domain of the SELFIE framework proved a valuable means of unpicking the various benefits, barriers and facilitators of the integrated service. At *Micro* level these included increased engagement of families, improved referral to preventative care services, and personalised health care; at *Meso* level the benefits of collocation, collaborative working and community outreach were described. Finally at the *Macro* level, the lack of engagement of the local care system was observed as were improvements to the access and availability of health and social care.

## Specific findings

### Micro-

#### Caregiver involvement

It is acknowledged that the voices of service users should be heard when decisions are made about their or their child’s medical care [42, 43]. Staff in the SCZ described how they consciously took time to explore individual patients and their family’s context and concerns, to better understand needs, wishes and feelings and gain parental consent: a key element of Early Help and enshrined in the UK’s Care Act (2014)[44]. Existing evidence describes how exhibiting such an interest in, and understanding of, patients cultural background, primary language, and cultural and faith practices, can help establish trust in the service, particularly amongst those communities previously suspicious of mainstream healthcare [10, 45–48].

#### Pro-active/preventative health

The aim of policymakers everywhere is to implement primary care that supports communities to better manage the factors influencing their health [49, 50]. However, exerting this control is inhibited in underserved populations by institutional, societal, and environmental barriers [51] this included GPs reporting a lack of resource for delivering preventative care [52, 53]. Despite these issues, the promotion of preventative care was reported as a successful element of the SCZ where staff described how their increased understanding of patients meant they could directly refer them to the most appropriate programmes.

Oral health was another key offer of the SCZ and is particularly important in the National Health Service where many children struggle to access the dental care they need [54]. The effectiveness of providing community routes of access to oral health care has been recognised previously [55], particularly amongst CYP [56], and the SCZ was able to link them to oral care through their partners in neighbourhood schools, another recognised route to accessing oral health care [57].

#### Tailored/personalised care

Clinicians described the benefits of taking additional time to understand both the individual but also their familial and community context. Understanding and accommodating these cultural needs, preferences, and broader social and cultural values of patients is an integral element of successful personalised care [43][58, 59]. The focus on developing a trusting and collaborative relationship with the patient (and family) displayed by clinicians in the SCZ is also known to improve compliance and clinical outcomes [60, 61].

### Meso-

#### Structural and organisational integration

Staff described how the health and social support components of the SCZ meant they were better able to address both clinical need and the various social determinants of health, as witnessed in similar service offers elsewhere [50, 62–64]. Such multi-disciplinary care offers have also previously supported an increase in inter-agency referrals [13], improved food security [14], access to care and patient experience, and greater confidence and trust from CYP and their families [15, 30, 65, 66].

School-based outreach appeared a particularly useful means of the SCZ engaging with more marginalised families where the use of informal settings and discussions recommended by NHSE for empowering disenfranchised patients [67]. The Department for Education (UK) has also flagged the importance of schools being used as a key means of engaging families and communities in the health and well-being of their children [68–71]. The potential of schools to serve youth in low-income, underserved communities is recognised internationally [72] and schools are regularly used in the United States and other high-income countries to engage families and provide support for mental health, chronic conditions, and preventative healthcare [41, 73–77]. Similarly in the UK there has been a renewed push to strengthen place-based partnership working with local communities as part of the integrational reforms associated with the 2022 Health and Care Act [78, 79]. Participants described how working in localised facilities enabled a better understanding and relationship with community groups active in their area, helping foster trust amongst the local population [79, 82, 83].

Staff described the benefits to teamwork, mutual respect and motivation, and professional development of sharing a space with colleagues. This echoes previous evidence that suggests collocating multi-disciplinary teams promotes efficient teamwork and collaborative communication [84–87] recognition of other professionals’ skills and contribution [62, 88] and reinforce shared beliefs and values [89].

### Macro-

#### Policies to integrate care

The NHS Health and Care Act of 2022 and a myriad of policies in advance of that were developed to deliver a more unified health and social care service capable of addressing the social determinants of health [8]. In the UK the introduction of integrated care systems were intended to drive true integration across settings however, participants reported a lack of ownership and leadership by the local integrated care system [90]. Not only in Birmingham but nationally, health and social care systems have been under immense financial pressure, with the resulting lack of capacity precluding prolonged institutional support of the SCZ service. This reluctance of the regional system to actively engage with integrated health and social care has been witnessed previously in the UK and looks set to continue in the absence of concerted evidence and consistent funding [12, 63, 91].

#### Streamlining referral processes

Typically referral process have proven particularly challenging for underserved populations due to a range of personal, and community-level factors including challenges of health literacy, and pronounced issues of competing life priorities, such as work, chronic disease and caring responsibilities [92–94]. With the understanding that streamlined referrals and those associated with a shorter follow-up period significantly improve connection rates [42, 94], including for CYP [95] there have been calls to improve the efficiency of pathways in underserved populations [96, 97]. The SCZ’s colocation provided further evidence of how attendance of CYP can be improved by enabling same day/same location referrals.

#### Translation/interpreter services

Staff delivering the SCZ described the benefits of a multilingual workforce and the use of interpreter services to meet the needs of their culturally diverse patients. The value and impact of staff and interpreters to improve the care and outcomes for culturally and linguistically diverse populations has been widely recognised [98, 99] including for children [100]. In the UK the NHS has released guidance for the commission of interpreting and translation services in primary care and its clear it has central role in place-based care [101, 102].

#### Strengths and limitations

Our rich dataset has provided valuable insight into the delivery of one of the UK’s first collocated place-based, integrated health and social support service. Participants were representative of the organisations involved in delivering the service and though their number (n=14) was lower than anticipated, the majority of active staff were interviewed, highlighting the issues in recruiting and funding staff in the early phases of the SCZ. The SELFIE framework proved a valuable tool in unpicking the experiences of delivering a collocated cross-sector community-based service [38, 39]. The validity of the findings was supported by regularly sharing and discussing the outputs of the analysis across the team [103]. Not every element of the SELFIE’s ‘Service Delivery’ domain were identified in our data set though all of our data was accommodated within it. We acknowledge that only gathering the experience of staff participants limits our understanding and in the next phase of the work we will interview service users including both CYP and their families.

## Conclusions

Integrated health and social care is seen as key to the future delivery of equitable health in the UK and beyond. This in-depth exploration of the SCZ pilot service offers further evidence of the benefits of a collocating multi-disciplinary staff in a single place-based service that delivers social support, health care, and preventative health. However, for it to be sustained greater system-level commitment, and leadership are needed.

## Data Availability

All data produced in the present study are available upon reasonable request to the authors

